# A Plasma Proteomic Ageing Clock Reflects Advanced Ageing in People with Untreated HIV and its Reduction Under Antiretroviral Therapy

**DOI:** 10.64898/2026.03.24.26348875

**Authors:** Barry Ryan, Mariam Ait Oumelloul, Sarah Rouached, Anna D. Juillerat, Laura Giacchetto, Christian W. Thorball, Isabella C. Schoepf, Jose R. Arribas, Berta Rodés Soldevila, Neeltje Kootstra, Peter Reiss, David Jackson-Perry, David Hans-Ulrich Haerry, Huldrych F. Günthard, Lena Bartl, Cédric Dollé, Doris Russenberger, Paolo Nanni, Tobias Kockmann, Marcel Stöckle, Luigia Elzi, Patrick Schmid, Alexandra Calmy, Daniel E. Kaufmann, Matthias Cavassini, Anders Boyd, Jacques Fellay, Johannes Nemeth, Philip E. Tarr

## Abstract

**Background:** Advanced ageing has been associated with an increased risk of serious disease endpoints in people with HIV (PWH). We conducted a longitudinal analysis to assess advanced proteomic ageing during untreated HIV infection and the effect of antiretroviral therapy (ART) on it by comparing the plasma proteome before and after ART initiation.

**Methods:** 416 protein abundance estimates were used to train a linear regression model predicting chronological age on 727 samples from Swiss HIV Cohort Study (SHCS) participants on long-term suppressive ART (median ART duration, 11.7 years). Advanced ageing was defined as age predicted by the proteomic ageing clock (PAC) minus chronological age. We evaluated the effect of successful ART on advanced proteomic ageing in an independent set of 80 PWH who had 4 longitudinal samples available, that is 2 samples during untreated HIV infection (>3 years apart, median interval between samples, 8·08 years (IQR 4·83–11·09)) and 2 samples during suppressive ART (>3 years apart, median interval between samples, 9·81 years (7·16–11·01)).

**Findings:** In the longitudinal test cohort, participants showed significantly higher proteomic age during untreated HIV infection than during suppressive ART, with a mean difference of 5.99 years (95% CI 4.25, 7.72), p = 0.0001. Thus, ART was associated with a marked reduction in proteomic advanced ageing. Although proteomic age remained higher than chronological age at all time points, linear interpolation of per-participant advanced ageing showed progressive normalisation towards chronological age during long-term suppressive ART. We validated these findings with our previously published epigenetic ageing study in the same cohort and extended those observations to the functional proteome, showing that proteomic data can capture acute immune signatures. Further, mediation analysis suggests that reversal of advanced ageing under ART is not driven by CD4+ or CD8+ T cell counts, indicating that the proteome captures ageing signals beyond immune reconstitution.

**Interpretations:** In a longitudinal study spanning more than 17 years, the advanced proteomic ageing observed during untreated HIV infection showed immediate and persistent deceleration under suppressive ART, demonstrating the importance of minimising the duration of untreated HIV infection.

**Funding:** Swiss HIV Cohort Study

**Research in context:** *Evidence before this study:* Current guidelines recommend prompt antiretroviral therapy (ART) initiation after HIV diagnosis, making it now difficult to quantify the potential effects of untreated HIV on advanced ageing. Biological ageing clocks serve as proxies for individual-level disease impact and are associated with serious disease endpoints in people with HIV (PWH). We searched PubMed for English-language reports from database inception to February 24, 2026, using combinations of the terms “HIV infection,” “antiretroviral therapy,” “proteomic ageing,” “proteomic clocks,” “proteomic advanced ageing,” and “age advancement.” We identified one study reporting that virally suppressed HIV infection is associated with a significant increase in proteomic ageing. We have previously shown in the well established longitudinal SHCS cohort with blood samples spanning >17 years and available both pre-ART and post-ART, that telomere length attrition and epigenetic ageing is accelerated during untreated HIV infection and that initiation of successful ART is associated with a significant reduction in accelerated ageing.

*Added value of this study:* To our knowledge, this is the first study to examine the impact of untreated HIV on the proteome using a proteomic ageing clock. Our results demonstrate that proteomic age is elevated before ART initiation and decreases significantly following successful viral suppression on ART. This reduction was not mediated by standard immunological markers (CD4+ and CD8+ T-cell counts,CD4:8 ratio). Compared with our previous epigenetics study, the proteome appears more responsive: advanced ageing increases more sharply during untreated HIV infection and is faster to decrease after ART initiation.

*Implications of all the available evidence:* Our findings demonstrate the importance of prompt ART initiation for PWH and reveal HIV-related ageing signals in the proteome that extend beyond immune reconstitution. Further, given the established association between advanced ageing and serious disease endpoints, this evidence motivates future studies into persistent advanced ageing to enable identification and stratification of high-risk PWH.

## 1. Introduction

Globally, approximately 40.8 million people live with HIV. HIV replication can effectively be controlled with modern antiretroviral therapy (ART), and timely access to and adherence to ART is now the key determinant of long-term health for people with HIV (PWH). Current ART guidelines recommend prompt initiation of therapy after HIV diagnosis; consequently, longitudinal follow-up in participants who have not begun ART is limited, making it challenging to assess both the health impact of untreated HIV and the benefits of ART within the same individuals ^1,2^. Here, leveraging a unique subset of participants from the Swiss HIV Cohort Study (SHCS) ^3^ who deferred ART for up to eight years and have banked plasma suitable for proteomic profiling, we quantify both the impact of untreated HIV infection and the effects of ART on biological ageing, as estimated by a plasma proteomic ageing clock (PAC).

Despite effective ART, PWH exhibit increased rates of age-related conditions such as coronary artery disease, obesity, and osteoporosis ^4–9^.This pattern aligns with evidence of advanced and accelerated ageing in PWH compared with the general population ^10–13^. Advanced ageing is defined as an elevated biological age compared to chronological age, and can serve a summary measure for the impact of illness on life expectancy and long-term quality of life ^14–16^. Biological age is typically estimated using algorithms trained on molecular markers, most commonly DNA methylation (referred to as epigenetic ageing clock (EAC)) and, more recently, proteomic profiles (referred to as PAC).

Advanced ageing in PWH has been linked to systemic inflammation and activation of pro-coagulatory pathways due to uncontrolled HIV viremia in the setting of untreated HIV infection ^17–19^. However, inflammatory biomarkers also remain persistently elevated in PWH despite effective ART, albeit to a lesser degree. ^11,12^. The molecular markers linking HIV viraemia to advanced ageing, and the extent to which ART modifies this process, remain incompletely understood. We previously showed in longitudinal analyses in participants of the SHCS that biomarkers of accelerated ageing that include telomere attrition and DNA methylation are markedly accelerated during almost 8 years of untreated HIV infection and that accelerated ageing is decelerated significantly and durably during almost 10 years of suppressive ART ^10,13^.

In this study, we extend these previous findings to the proteome. Proteomic measurements provide functional insight into the mechanisms and dynamics of advanced biological ageing. Furthermore, proteomic profiles tend to reflect disease effects more rapidly than epigenetic measurements ^20^, offering finer temporal resolution around HIV diagnosis and ART initiation than slower-moving epigenetic readouts. Although PACs are relatively new compared to EACs, they already capture advanced ageing and are associated with increased risk of prevalent age-related serious disease endpoints in PWH ^11,21^. In summary, in this study we have characterised the proteomic ageing dynamics of PWH enrolled in the SHCS, using longitudinal samples spanning more than eight years of untreated HIV infection and nearly ten years of suppressive ART, by quantifying the impact of untreated HIV infection and the effect of ART on proteomic advanced ageing.

## 2. Methods

### 2.1. Study design

The overall workflow is shown in Figure 1. The training cohort comprised PWH on long-term suppressive ART, as evidenced by an undetectable HIV viraemia at the time of sampling. The test cohort comprised the same participants, samples, and timepoints as in our previous longitudinal telomere length and epigenetic accelerated ageing studies, for whom detailed inclusion and exclusion criteria were previously reported ^10,13^. Briefly, all SHCS participants were eligible if they had 4 stored blood samples available, that is 2 samples (at least 3 years apart) during untreated HIV infection plus 2 samples (also at least 3 years apart) during suppressive ART.

**Figure 1.**
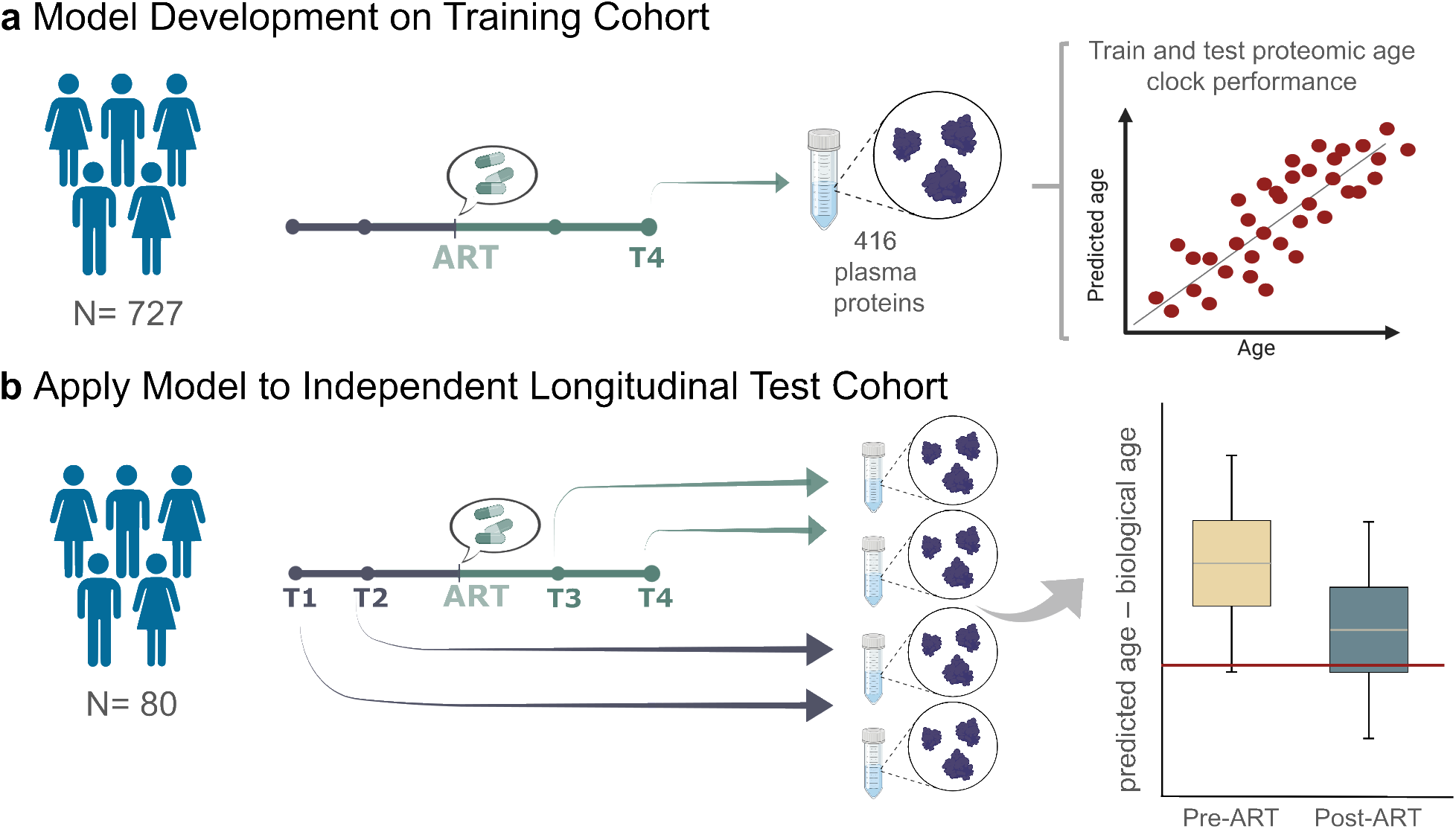
Proteomic ageing clock training and validation. 416 plasma proteins from 727 PWH under ART with suppressed HIV viraemia, were used to train a ridge regression model. This model was applied to the longitudinal cohort of PWH, highlighting the effect of untreated HIV infection and of effective ART on their biological age.

We trained a linear model to predict age in the training cohort and then applied it to the test cohort. Advanced ageing was defined as PAC-predicted age minus chronological age. We compared advanced ageing within the test cohort at each cross-sectional time point to assess whether there was an effect of untreated HIV infection and of suppressive ART on proteomic advanced ageing.

### 2.2. Study participants

All data was generated as part of the SHCS, a prospective observational cohort that has been following people living with HIV in Switzerland since 1988^3^. Population demographics for both cohorts are shown in Table 1. In the training cohort, proteins were measured at a single time point during effective ART, i.e. HIV viraemia (HIV RNA) was undetectable. Participants were excluded if they appeared in the test cohort or if they had detectable HIV viraemia at the time point closest to the date at which the sample was taken. Participants in this training set were used to train the PAC.

**Table 1:** Participant demographics, immunological cell counts and ART duration in the training cohort during suppressive ART and the test cohort at the final time point (T4)

|  | Training Cohort | Test Cohort (T4) |
| --- | --- | --- |
| **Demographics** |  |  |
| N Participants | 727 | 80 |
| Age, years: median [IQR approx] | 56.0 [49.5, 62.5] | 53.5 [49.0, 58.0] |
| Age, years: range | 30–91 | 38–74 |
| Male sex at birth | 635 (87.3%) | 52 (65.0%) |
| **Smoking Status** |  |  |
| Current | 228 (31.4%) | 45 (56.3%) |
| Past | 231 (31.7%) | 14 (17.5%) |
| Never | 268 (36.9%) | 21 (26.2%) |
| **Race/Ethnicity** |  |  |
| White | 630 (86.6%) | 76 (95.0%) |
| Black | 21 (2.9%) | 1 (1.25%) |
| Hispano-American | 3 (0.5%) | 1 (1.25%) |
| Asian | 7 (0.9%) | 2 (2.5%) |
| Unknown | 66 (9.1%) | 0 (0%) |
| Time Since HIV Diagnosis, years: mean ± SD | 14.4 ± 7.2 | 22.1 ± 4.6 |
| **Immune and Virological Measures** |  |  |
| HIV RNA viraemia mean ± SD | 0 ± 0 | 1.81 ± 9.28 |
| HIV RNA viraemia median[IQR approx] | 0 [0, 0] | 0 [0, 0] |
| CD4+ mean ± SD | 636.2 ± 98.39 | 719.90 ± 280.56 |
| CD4+ median [IQR approx] | 594.5 [424.8, 800.0] | 646.0 [541.5, 896.0] |
| CD8+ mean ± SD | 844.8 ± 408.98 | 758.20 ± 366.53 |
| CD8+ median [IQR approx] | 775.0 [549.5, 1057.5] | 689.5 [514.75, 920] |
| **Duration of ART** |  |  |
| Years since ART start: mean ± SD | 13.1 ± 6.7 | 11.02 ± 3.64 |
| Years since ART start: median [IQR approx] | 12.9 [7.7, 18.2] | 10.71 [8.55, 12.71] |

In the test cohort, proteomics were measured and proteomic ageing was calculated at time point 1 (T1; first available sample before ART start), time point 2 (T2; last available sample before ART start; >3 years after T1), time point 3 (T3; first sample at which HIV RNA was suppressed after ART start, defined as *<*100 copies per mL) and time point 4 (T4; most recent available sample during suppressive ART [defined as *<*100 copies per mL], >3 years after T3). 67 of the 80 participants had two samples available before starting ART (T1, T2) and two samples after starting ART (T3, T4), resulting in four samples per participant. 13 participants had only two samples post ART start (T3, T4). We obtained clinical, immunological, and HIV-related variables to evaluate their association with the rate of proteomic ageing in each study phase including chronological age; sex; smoking status (current, past, or never); HIV viraemia (maximum log10 HIV RNA); CD4+ T-cell count; CD8+ T-cell count and CD4:8 ratio.

We show the values at T4 in Table 1 for the test cohort, as this is the time point at which training and test cohort participants both have had suppressed HIV viraemia for a prolonged duration of time. As per Table 1, the training cohort had a higher proportion of males and are older than the test cohort. Models were adjusted for these characteristics as discussed in supplementary appendix A5. Differences in immunological, viral, and ART parameters are the main effects being tested longitudinally in the test cohort, and therefore were not adjusted for during modelling.

### 2.3. Proteomics procedures

We established a neat plasma proteomics workflow with automated sample preparation, rigorous batch, plate, and position quality control, standardised data acquisition, and applied it to two sub-cohorts within the SHCS. Proteins were measured via high-throughput mass spectrometry with quantification and performed using the data independent acquisition neural network software ^22^. A total of 1,456 proteins were detected across all training and test cohort samples. A total of 626 proteins passed processing procedures and were common across both training and test cohorts. Batch correction was performed across the training and test cohorts using ComBat to ensure generalisation of the model to the test set ^23^. See Appendix for full details.

### 2.4. Proteomic ageing clock modelling

A plasma PAC was established by training a series of linear models on the data of the training cohort. To mitigate confounding, we first adjusted each protein’s abundance for sex, study site, and genetic ancestry. Specifically, for each protein we fit an ordinary least squares (OLS) regression on sex, study site, and the first three genetic principal components (PCs), and retained the residuals as covariate-adjusted proteomic features for downstream PAC training.

We used ElasticNet regression for feature selection and we trained the PAC as a ridge regression model on the selected protein subset using the full training cohort. We performed 5-fold cross-validation to tune hyperparameters for ElasticNet and we employed a relaxed fit on the ridge regression model to reduce shrinkage-induced bias and improve generalisability to the test cohort.

Using the selected proteins, we performed pathway enrichment analyses, weighting features by their coefficients from the ridge regression model. We then constructed a protein-protein interaction (PPI) network with the STRING ^24^ database, clustered the network, and conducted additional enrichment on the resulting clusters to provide a more fine-grained functional characterisation.

### 2.5. Statistical analyses

Statistical analysis was performed in the independent, held-out test cohort at four cross-sectional time points, with duration of ART, in years, at each time point shown in Figure A4. We tested whether suppressive ART is associated with a change in advanced ageing, defined as the difference between biological age (as predicted by the PAC) minus chronological age. We performed OLS inference using the Statsmodels package in python, with participant-clustered robust standard errors, to assess if there is a statistically significant change in advanced ageing once HIV viraemia has successfully been suppressed during ART ^25^. We report estimated mean differences in advanced ageing versus the four reference time points, with 95% confidence intervals and two-sided p-values. We further compared our previous epigenetic accelerated ageing findings ^10^ with PAC results at each of the four time points to assess overall concordance. Mediation analyses were performed using the R package mediation ^26^. Advanced ageing was used as the outcome, and the exposure was time since ART initiation, defined continuously so that negative values indicate pre-ART samples and positive values indicate on-ART samples, as per Figure A4. To assess whether immune recovery mediates the association between time since ART and advanced ageing, we evaluated candidate mediators measured at cross-sectional time points T2, T3, and T4. The candidate mediators were plasma HIV viraemia, CD4+ T-cell count, CD8+ T-cell count, and the CD4:CD8 ratio. All mediation models were adjusted for sex, study site and ethnicity.

## 3. Results

### 3.1. PAC evaluation

Figure 2 summarises PAC metrics in the training and test cohorts. Panel a shows that principal component analysis (PCA) across time points demonstrates high between-cohort variation at earlier (pre-ART) time points, with clear separation along the second principal component. At later, post-ART time points, the training and test batches overlap more closely, indicating improved biological concordance.

**Figure 2.**
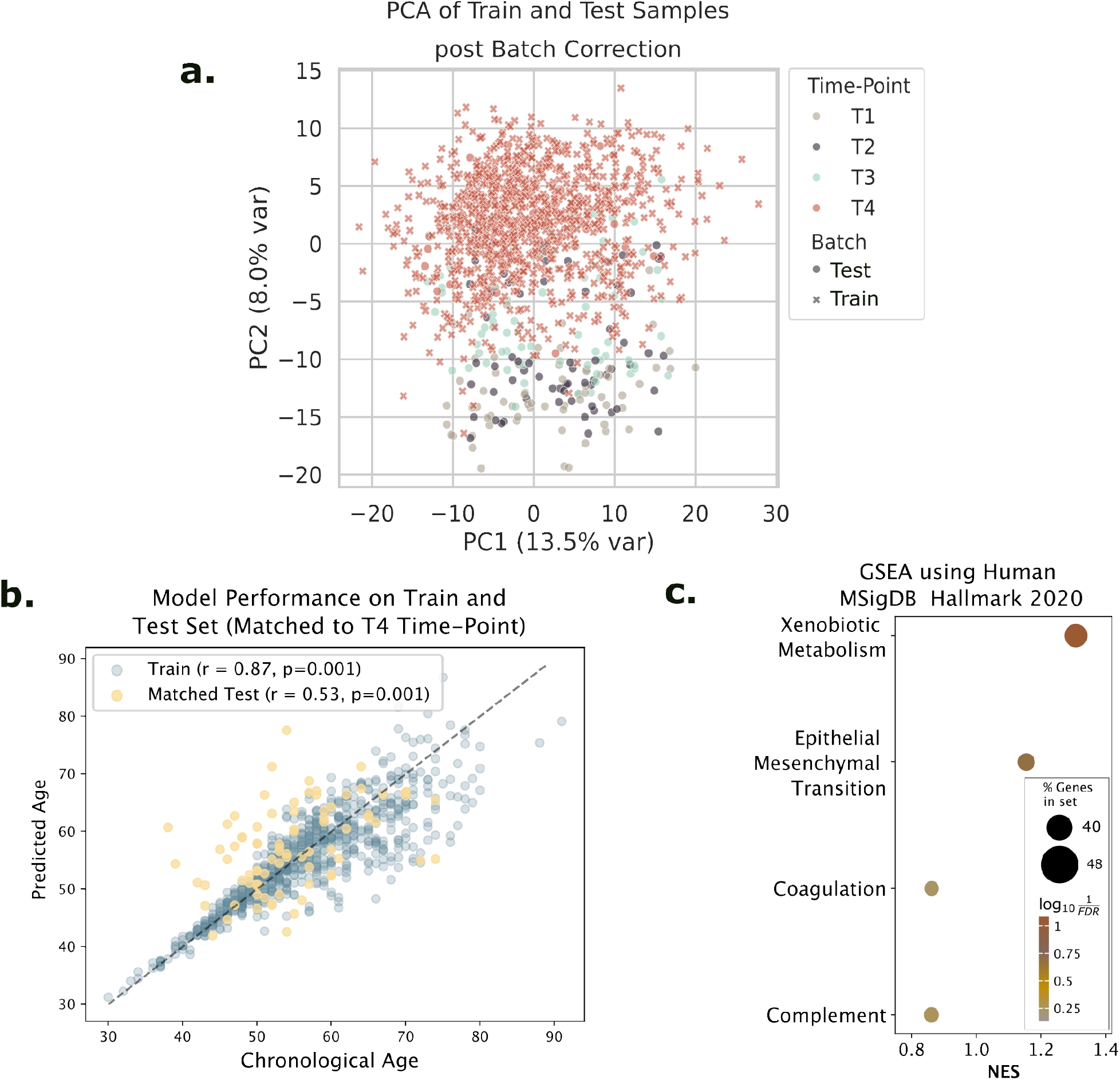
Proteomic ageing clock training metrics; a, principal component analysis of early time points reveals variance across batches. b, Proteomic ageing clock model performance in the training cohort and in a matched test cohort (participants at the T4 time point). c, gene set enrichment analysis (GSEA) highlights pathways associated with PAC-estimated age

Panel b shows residuals for the training and test cohorts, restricted to the T4 time point to match the biological conditions of the training cohort. After ElasticNet–based feature selection, 416 proteins were retained and used to fit a ridge-regression PAC model. In the training cohort, the model achieved *r* = 0.87 and a mean absolute error (MAE) of 3.34 years. As shown in Table A1, MAE was lower at younger ages, consistent with the age-weighted loss and distribution matching. In the test cohort at T4, performance was lower (*r* = 0.53 and MAE 6.62 years). Such differences are expected, with good overall concordance between cohorts and no evidence of systematic bias in the test cohort at T4.

As shown in Table A2, at earlier time points, and therefore younger ages, the MAE is higher in the test cohort, with predictions biased towards older ages. All participants in the training cohort were on suppressive ART. In comparison, those in the test cohort had not received ART at time points T1 and T2, and at subsequent time points (T3 and T4) had successfully suppressed HIV viraemia. Given the high concordance at T4 and mitigation measures of age-weighted loss and matched age distributions, we attribute this systematic increase in MAE to biological variation associated with pre-ART status at the earlier time points.

Using ridge-regression coefficients as protein weights, we performed pathway enrichment and network analyses to characterise the functions of the most informative proteins. Both approaches implicated coagulation, complement, and xenobiotic metabolism as key contributors to ageing signals in the training cohort. Coagulation and complement pathways are immune pathways well recognised in the biology of ageing and are enriched for proteins recently reported to be dysregulated between so called viraemic and HIV elite controllers ^27^. Xenobiotic metabolism is a pathway which converts drugs into their active form, and therefore, it should be a strong indicator of ART impact.

### 3.2. Advanced ageing during untreated HIV and its reduction under suppressive ART

The results presented in Figure 3a show increased advanced ageing during untreated HIV infection and its marked reduction once HIV viremia is suppressed during ART. As per Table A3, during untreated HIV infection there is significantly higher proteomic advanced ageing compared to samples from the same individuals after HIV viremia is suppressed, with a mean difference of 5.99 years (95% CI 4.25, 7.72), p = 0.0001. Interestingly, there is a statistically significant drop in advanced ageing already at T3 (Figure 3a and Table A4), that is at the first time point that HIV viremia is suppressed, with T3 occurring a median 1.55 years (IQR, 0.68-1.64 years) after ART start. Although the average proteomic age remains higher than chronological age at all time points, linear interpolation per-participant estimates that advanced age will converge to chronological age 16.40 years (95% CI 13.55, 19.25) after ART start (Supplementary File 1). These findings were robust to adjustment for technical covariates and consistent across repeated post-ART measurements.

**Figure 3.**
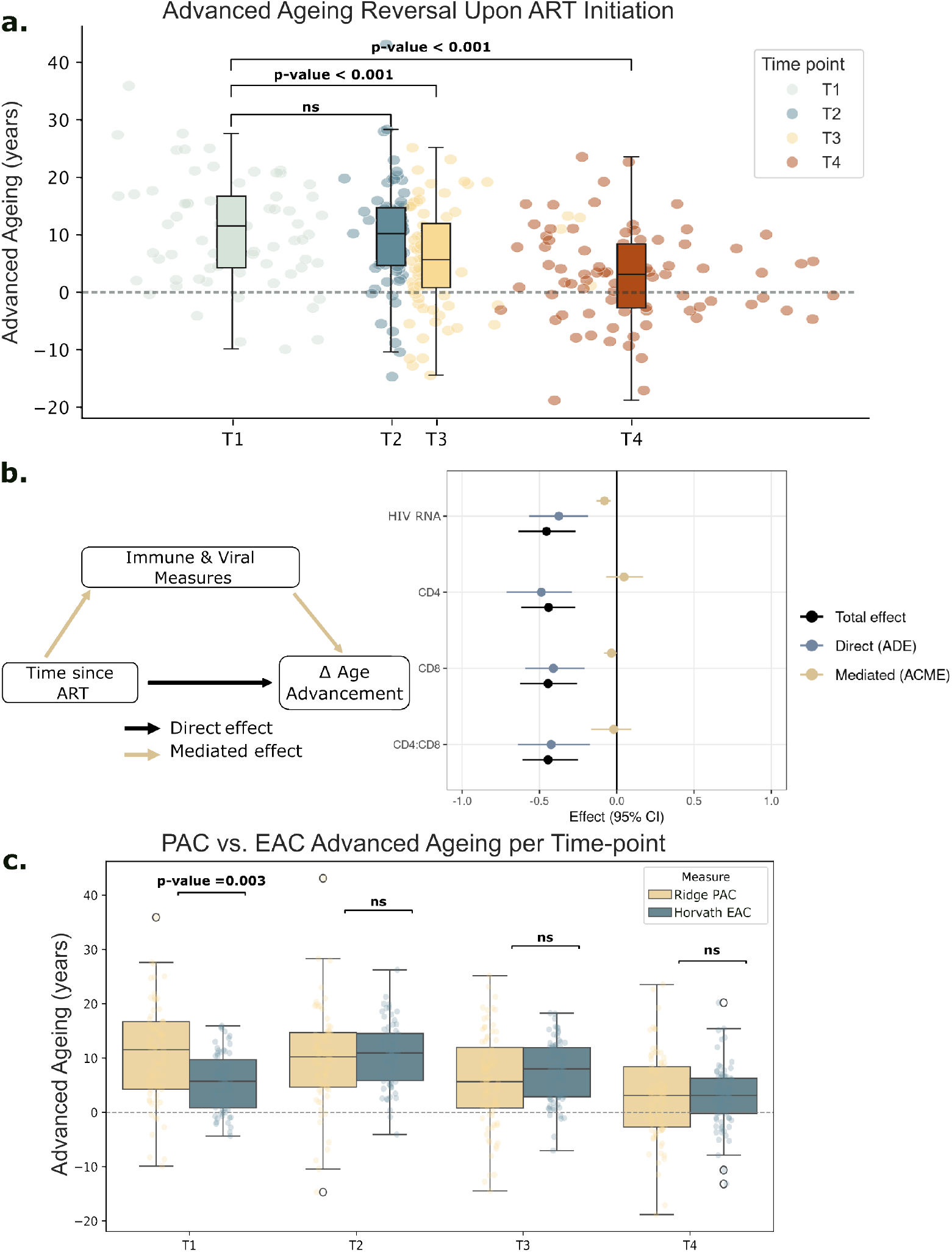
Evidence of advanced ageing reversal under ART; a, Cross-sectional analyses show an immediate reduction in advanced ageing at ART initiation, and a convergence towards chronological age under long-term ART. b, Mediation analysis shows that while advanced ageing reversal is driven by a reduction in HIV viraemia, it was not found to be mediated by conventional immune CD4+ and CD8+ T-cell counts or CD4:8 ratio. c, Cross-sectional comparison between proteomic ageing clock (PAC) and Horvath epigenetic ageing clock (EAC) at T1 shows a statistically significant difference, likely reflecting the immediate impact of untreated infection in the ageing process captured by the proteomic ageing clock (PAC) compared to the epigenetic ageing clock (EAC).

### 3.3. Mediation analysis

As shown in Figure 3b, mediation analyses, adjusted for sex and ethnicity, showed a significant indirect effect of ART on advanced ageing through plasma HIV RNA viraemia (ACME *β* = -0.079, 95% CI -0.132 to -0.041; p-value<0.001). This result indicates that reductions in HIV viraemia partially mediate the decrease in advanced ageing in PWH, highlighting the efficacy of ART and the importance of prompt, sustained virological suppression.

In contrast, our results indicate that neither CD4-count (ACME *β* = 0.047, 95% CI*−* 0.073 to 0.174; p-value=0.41), nor CD8-count (ACME *β* = *−* 0.034, 95% CI *−*0.082 to 0.001; p-value=0.054), or the CD4+:CD8+ ratio (ACME *β*= *−*0.020, 95% CI*−* 0.154 to 0.099; p-value=0.73) significantly mediate the reversal of proteomic advanced ageing in PWH once HIV viraemia is suppressed following the initiation of ART. This suggests that immune reconstitution, as captured by CD4 and CD8 cell counts, does not mediate deceleration of proteomic ageing and that the proteome reflects ageing signals beyond these immunological measures.

### 3.4. Comparison of proteomic and epigenetic ageing

Figure A5 shows moderate correlation (r=0.44, p-value=0.0001) between the PAC, reported on here, and the EAC predictions on the longitudinal test cohort we previously reported ^10^. This suggests that both PAC and EAC capture shared underlying patterns of biological ageing in PWH. Interestingly, as the PAC was trained on a cohort of participants with HIV, compared to the EAC which was not, it supports the findings from mediation analysis that reversal of biological ageing is not driven solely by immunological status.

Figure 3c shows shared patterns of advanced ageing reduction in both PAC and EAC. At T1, the first available sample in each participant, PAC indicates significantly greater advanced ageing than EAC (Table A7), consistent with PAC capturing immediate biological changes during untreated HIV infection that EAC may be slower to detect. The opposite pattern is seen at T3, where the PAC shows a slightly lower mean advanced ageing than the EAC, although this difference does not reach statistical significance (p-value = 0.066). At each time point, PAC has a larger standard deviation than EAC, likely due to the smaller PAC training set (n=727) compared with the Horvath EAC (n=8000) ^28^.

## 4. Discussion

Advanced ageing has been associated with HIV infection and linked to increased serious disease endpoints in PWH. We trained a PAC using samples from the SHCS and used it to assess the effect of untreated HIV infection and of successful ART on advanced ageing in an independent longitudinal subset of SHCS participants with samples spanning over 8 years of untreated infection and nearly 10 years of suppressive ART. We observed advanced ageing during untreated HIV infection and its statistically significant reduction after successful suppression of HIV viraemia was attained by means of ART. Further, linear interpolation of per-participant advanced ageing showed progressive normalisation towards chronological age during long-term (almost 10 years) of suppressive ART. Our findings corroborate and extend prior epigenetic studies that also report reversal of advanced ageing under ART ^10,18,19,29^.

PACs have previously been shown to capture advanced ageing in HIV ^11,21^. Our PAC was trained on samples from PWH with undetectable viraemia and applied to a longitudinal cohort spanning time points before and after ART initiation. As the PAC is a linear combination of plasma protein abundances, with both the training and test cohorts comprised of PWH, the between-group differences by HIV status are minimised and the model primarily captures biological variation unrelated to HIV status. In this longitudinal setting, the principal time-varying exposure is ART initiation. Analyses at four time points before and after ART start show a marked and statistically significant reduction of advanced ageing under successful ART. Interpretability analyses highlighted that the most influential proteins map to complement and coagulation inflammatory pathways, which have established links to advanced ageing ^30,31^. Consequently, our finding suggests that reduced advanced ageing under ART is associated with changes in these pathways.

Mediation analysis extends these findings. Reduction of advanced ageing was not found to be mediated by conventional HIV recovery markers of CD4+ or CD8+ T cell counts in this study. Instead, mediation analysis indicates that reductions in advanced ageing during suppressive ART are mediated in part by reductions in HIV viral load. Following the interpretability findings, this could suggest the involvement of alternative immunological and inflammatory pathways. Overall, these findings help explain why some PWH continue to experience poor health outcomes on ART and suggest that optimising long-term care will require addressing processes beyond ART’s direct effects on immune reconstitution as measured by CD4+ and CD8+ counts alone ^12^.

PAC predictions correlated moderately with EAC ages in the same participants, indicating that proteomic and epigenetic clocks capture shared biological variation and validating the PAC model. Both the PAC and the EAC show significantly reduced advanced ageing under ART, which is also consistent with the observed attenuation of telomere shortening in the same longitudinal samples ^13^. Taken together, these findings support long-term biological benefits of ART in PWH. Notably, the PAC showed a statistically significant larger mean advanced ageing at T1, compared to the EAC. This difference could be explained by modality-specific dynamics of each clock. Epigenetic measures operate at the chromatin level and tend to integrate signals over longer time scales, potentially missing immediate responses to primary HIV infection in this context ^20^. Conversely, proteomic profiles can change rapidly with acute inflammatory and immunological perturbations, making PACs more temporally responsive, consistent with our finding at T1.

The magnitude and clinical interpretation of differences between proteomic and epigenetic measures of advanced ageing in PWH needs to be independently replicated before considered a true finding. In this study, we use the Horvath epigenetic ageing clock because it is trained on only 353 CpG sites, making it straightforward to interpret and compare across datasets ^28^. Future work should systematically compare epigenetic and proteomic clocks, as findings will be sensitive to the specific proteins and CpG sites used to train each model.

Future work should replicate this study using a PAC trained in an HIV-negative cohort. PACs often exhibit limited generalisability and sensitivity across studies because of differences in proteomic platforms, preprocessing, participant populations, statistical methods, and sampled tissues or cell types ^32^. In this study, reusing previously published PACs was inhibited by incomplete overlaps in measured proteins. Further, even where overlaps are perfect, platform and modelling differences can distort predictions. Training the PAC within a HIV cohort using a linear model may absorb HIV-related proteomic shifts into the baseline, therefore reducing HIV related signal. Conversely, an external PAC not trained on PWH would provide an independent reference which would capture the joint impact of HIV and ART on biological ageing. Given the prior evidence that HIV infection, despite suppressive ART, is associated with advanced ageing in comparison to participants without HIV ^11^, such an approach would likely show higher advanced ageing than what is reported here.

Importantly, subsequent research should link the proteomic changes observed here to hard clinical outcomes, such as cardiovascular and other events. As the SHCS began systematic clinical endpoint capture only starting in 2000 and the number of potential endpoints is large relative to our sample size, the current study is underpowered for enrichment or outcome-association analyses, as previously published ^10^. Further, although our findings are consistent with previous studies and align with the known natural history of HIV, our results are limited by sample size which reduces the precision and power of inferences and mediation analyses, therefore conclusions should be drawn with caution.

In conclusion, we have developed a plasma PAC for PWH and demonstrated that proteomic ageing is advanced during untreated HIV infection and that proteomic age advancement is reduced with suppressive ART. The approx 6-year reduction in proteomic age post-ART highlights the reversibility of advanced ageing based on plasma proteome signatures and supports its use as a sensitive, clinically interpretable endpoint for monitoring recovery and evaluating adjunctive interventions. Mediation analyses indicate that the reduction in advanced ageing is not driven by CD4+ or CD8+ cell counts, suggesting that the proteome captures ageing signals beyond immune reconstitution. Finally, we validate our findings with prior ageing studies, in particular, demonstrating the benefit of extending epigenetic clock observations to the functional proteome.

## Data Availability

Deidentified individual participant data used in the study can be made available for investigators upon request to the corresponding author.
The proteomics data has been deposited to the ProteomeXchange Consortium via the PRIDE32 partner repository with the dataset identifier PXD075234. This data can also be made available for investigators upon request to the corresponding author.

## 5. Contributors

SR, AJ, PT, and JN contributed to study design. MS, AC, MC, LE, PS, GW, PT contributed to patient recruitment. LG, PN, TK and PT contributed to data acquisition. BR, MAO, TK, and JN analysed the data. BR, MAO, PT, JF, and JN verified the underlying data and drafted the manuscript. All authors contributed to critical review and revision of the manuscript. PT, JF, and JN had final responsibility for the decision to submit for publication.

## 6. Data sharing

Deidentified individual participant data used in the study can be made available for investigators upon request to the corresponding author.

The proteomics data has been deposited to the ProteomeXchange Consortium via the PRIDE ^33^ partner repository with the dataset identifier PXD075234. This data can also be made available for investigators upon request to the corresponding author.

## 7. Acknowledgements

This study was financially supported by the Swiss HIV Cohort Study (SHCS; project 918). SHCS data are gathered by the five Swiss university hospitals, two Cantonal hospitals, 15 affiliated hospitals and 36 private physicians (listed in http://www.shcs.ch/180-health-care-providers). The authors acknowledge the effort and commitment of SHCS participants, investigators, study nurses, laboratory personnel, and administrative assistance by the SHCS coordination and data centre.

