## Appendix for "A Plasma Proteomic Ageing Clock Reflects Advanced Ageing in People with Untreated HIV and its Reduction Under Antiretroviral Therapy"

#### 1 Urea-assisted digestion of plasma proteins

For each sample, 5 µl of human plasma was transferred to a 96-well PCR plate (Eppendorf twin.tec) prefilled with 55 µl of 8 M urea in 50 mM Triethylammoniumbicarbonat (TEAB). Samples were sonicated (sonication bath) for 5 min prior to reduction with 5 mM tris(2-carboxyethyl)phosphine (TCEP) and alkylation with 15 mM chloroacetamide (ClAA) at 30°C for 30 min.

The protein digestion protocol was automated on an Opentrons Flex robot (Opentrons) equipped with a Flex 1-Channel 50 µL, a Flex 8-Channel 50 µL pipette head, a Flex Gripper, a Temperature Module GEN2 carrying an Opentrons 24 Well aluminum Block, and a Heater-Shaker Module GEN1 carrying an Opentrons 96 PCR Heater-Shaker Adapter. In short, 35 µl of the above described plasma sample was diluted by adding 185 µl of 50 mM TEAB, pH 8.5 to a final urea concentration of 1.2 M, reduced and alkylated by incubation with TCEP and ClAA for 30 min at 37°C and shaking at 600 rpm, and finally digested overnight by off-deck incubation at 37°C after adding 1 µg of sequencing grade modified trypsin (Promega). Digestion was stopped by adding 15 µl of 10% trifluoroacetic acid (TFA). During protocol execution trypsin stock solution (0.4 ng/µl) was stored in the aluminum block cooled to 8°C, while CIAA (500 mM in ddH2O), and TCEP (100 mM in ddH2O) were stored at RT in a NEST 12 Well Reservoir 15 mL. The Opentrons .json file is provided as supplementary material and can be opened using the Opentrons Protocol Designer. For each sample an amount equivalent to 0.3 absorbance units (280 nm) was transferred onto single-use StageTips (Evotip Pure, Evosep Biosystems) according to the manufacturer's instructions including iRT peptides (Biognosys).

#### 2 Liquid chromatography mass spectrometry

LC-MS analysis was performed on a timsTOF flex (Bruker) equipped with a CaptiveSpray 2 ion source (Bruker), and directly coupled to an Evosep One (Evosep Biosystems) chromatography unit. Peptide mixtures were separated on a PepSep C18, 8 cm x 150 µm, 1.5 µm column (PN 1893470) kept at 40°C using the 60 samples-per-day method. The mass spectrometer was operated in dia-PASEF mode [1]. TIMS ramps were acquired in positive polarity from 100 to 1700 m/z and inverse mobilities (1/K0) from 0.85 to 1.30 V*s/cm2 with a synchronized ion accumulation and ramp time of 100 ms. Each MS ramp was followed by 8 MS/MS ramps covering the mass range of 475 to 1000 m/z and the mobility range of 0.85 to 1.27 V*s/cm2 in 21 quadrupole isolation windows of 25 Da (estimated cycle time 0.95 s). During MS/MS scans collisional energy was ramped linearly from 20 to 59 eV across the mobility range of 0.6 to 1.6 V*s/cm2.

#### 3 LC-MS data management and availability

LC-MS data was handled using the laboratory information management system b-fabric [1] and all relevant data have been deposited to the ProteomeXchange Consortium via the PRIDE (http://www.ebi.ac.uk/pride) partner repository with the data set identifier PXD075234 [2-4].

### 4 Protein identification and quantification

The recorded LC-MS data was processed using DIA-NN 1.8.2 beta 8 using a predicted spectral library (aka library-free mode) and match-between-runs (MBR) [5]. In short, chromatographic features were assigned to the canonical human reference proteome (UP000005640) obtained from UniprotKB (2024/10/15), after in-silico digestion to fully tryptic peptides including a max. of 1 missed cleavage (min. peptide length: 6 aa; max. peptide length of 30 aa), assuming fixed carbamidomethylation of Cysteine (Unimod#4) and variable Methionine oxidation (Unimod#35), taking into account precursor charge states +2 and +3. Mass accuracy was fixed to 15 ppm for MS and MS/MS level analysis. Protein (group) abundances were estimated using the maxLFQ method and reported at 1

### 5 Proteomics Processing

Proteins in plasma serum were quantified by liquid chromatography mass spectrometry (LCMS) in two separate runs, each using a different reference. The first reference assigned each peptide to a single protein only, whereas the second allowed peptides to map to alternative proteins. Protein intensities from both runs were log2-normalized. To correct for run- and instrument-level batch effects, samples were normalized using bridge (master) samples; these bridge samples were then removed. The two runs were merged to maximize protein coverage, using values from one run to fill missing (NA) values in the other when available. Remaining missing values were imputed with the minProb method for proteins with less than 50% missingness. Batch correction for plate effects was performed within each cohort. We then restricted analyses to proteins present in both cohorts and applied an additional batch correction to align the training and test sets, as described below.

### 6 Batch Correction using ComBat

Batch correction was performed using the ComBat, an empirical Bayes method for batch effect correction in high-throughput omics data (e.g. microarrays, RNA-seq, DNA methylation, and proteomics). It removes unwanted variation due to technical batches (e.g., different plates, runs, labs) while aiming to preserve biological signal.

As the ART status varied over time in our longitudinal test cohort, we estimated ComBat batch-effect parameters using only samples from T4, which best matched the biological state of the training cohort (median time under ART of 11.7 years with RNA viral load of 0 copies/ml) To preserve ART-related biological variation, we then froze these parameters and applied ComBat to each time-point in the test cohort using the same estimates. The ComBat-adjusted values were subsequently used in downstream analyses.

### 7 Elastic Net Model

Elastic Net is a regularised linear model that blends L1 (lasso) and L2 (ridge) penalties, enabling both variable selection and coefficient shrinkage. It handles groups of correlated predictors better than lasso alone, often keeping or dropping them together. The mixing parameter $\alpha$ controls the L1–L2 trade-off, while $\lambda$ sets the overall regularisation strength; predictors are typically standardised so penalties act uniformly. Models are commonly fit via coordinate descent, with $\alpha$ and $\lambda$ tuned by cross-validation. The objective function is:

$$\hat{\beta} = \underset{\beta}{\operatorname{argmin}} \left( \sum_{i=1}^{n} (y_i - \beta_0 - \sum_{j=1}^{p} x_{ij}\beta_j)^2 + \lambda_1 \sum_{j=1}^{p} |\beta_j| + \lambda_2 \sum_{j=1}^{p} \beta_j^2 \right) \tag{1}$$

Figure A1 shows a strong negative trend in the ElasticNet residuals, indicating underfitting consistent with shrinkage bias. This bias must be addressed given the age distribution shift between cohorts. If uncorrected, it could introduce estimation bias for younger participants that is unrelated to ART. Resultingly, to eliminate this effect, we used ElasticNet model for feature selection only.

#### 8 Ridge Regression Model

Ridge regression is a linear model with L2 regularisation that shrinks coefficients toward zero to reduce variance and improve generalisation, especially under multicollinearity. It typically retains all predictors (no hard variable selection), shrinking correlated coefficients together. The regularisation strength $\lambda$ controls the amount of shrinkage; predictors are usually standardised so the penalty acts uniformly, and the intercept is left unpenalised. The model has a closed-form solution via $(X^T X + n\lambda I)^{-1} X^T y$, and $\lambda$ is commonly chosen by cross-validation. The objective can be written as:

$$\hat{\beta}_{ridge} = \underset{\beta}{\operatorname{argmin}} \left( \sum_{i=1}^{n} \left( y_i - \beta_0 - \sum_{j=1}^{p} x_{ij}\beta_j \right)^2 + \lambda \sum_{j=1}^{p} \beta_j^2 \right) \tag{2}$$

The PAC model trained with ridge regression fits the data better than the ElasticNet model. Ridge regression on a subset of proteins largely mitigated shrinkage bias and yielded a better overall fit, as evidenced by residuals that are closer to normally distributed in Figure A2.

#### 9 Proteomic Age Clock Training Considerations

Proteomic ageing clocks commonly exhibit age-dependent error, with larger absolute errors at younger ages. To mitigate this bias, we applied age-based loss re-weighting. Within 10-year chronological-age bins, we computed inverse-frequency sample weights so that each bin contributed equally to the mean absolute error. As the training and test cohorts differ in age distributions, as per Table 1 and Figure A1, we additionally trained models with importance weights to approximate a target age distribution centred at 45 years (SD 10 years), with the aim of reducing mean absolute error among younger participants.

To limit shrinkage-induced bias and accommodate systematic differences between cohorts, the ElasticNet model was used solely to identify informative proteins (Figure A2). We performed 5-fold cross-validation to tune the L1:L2 mixing parameter and the overall penalty, refit ElasticNet on the full training set with the optimal values, and retained features with non-zero coefficients. These features were then carried forward into the final ridge regression model.

#### 10 Sensitivity Analysis of PAC

We performed sensitivity analyses of the PAC within the training cohort to assess whether predictions were influenced by covariates (e.g., sex, sampling centre), with results summarised in Table A5. Although we residualised the data for sex, sampling centre, and genetic PCs, some covariates remain nominally significant. This likely reflects residual structure rather than true model sensitivity. Linear residualisation removes only main linear effects; non-linear relationships, interactions (e.g., covariate $\times$ time), or batch-specific variance differences can leave detectable associations. In addition, unequal representation across centres and sexes increases statistical power in the largest groups, so very small residual effects can appear significant. For example, larger centres contribute more samples and thus more weight in hypothesis tests. Consistent with this interpretation, effect sizes are small, and re-estimating models that explicitly adjust for centre and sex (or using centre-cluster-robust standard errors or mixed-effects with random centre intercepts) would not materially change the primary results.

As the training cohort was originally recruited for a cardiac screening study with an equal 50:50 split of cardiac cases and controls, we also tested for sensitivity to case status. As expected from the balanced design, and reported in Table A6, we found no evidence that case status influenced PAC predictions (p-value=0.588).

#### 11 Supplementary Figures

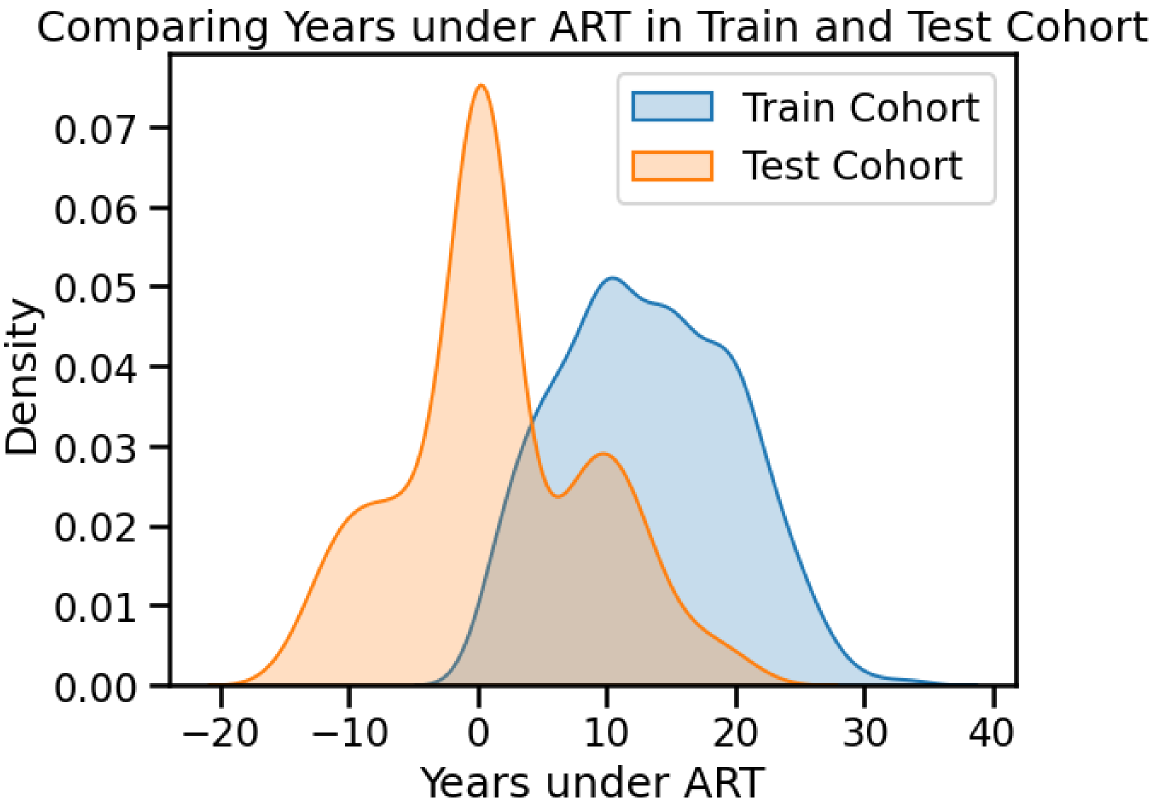


Figure 1: Age distributions of training and test cohorts. All time points were used for the test cohort.

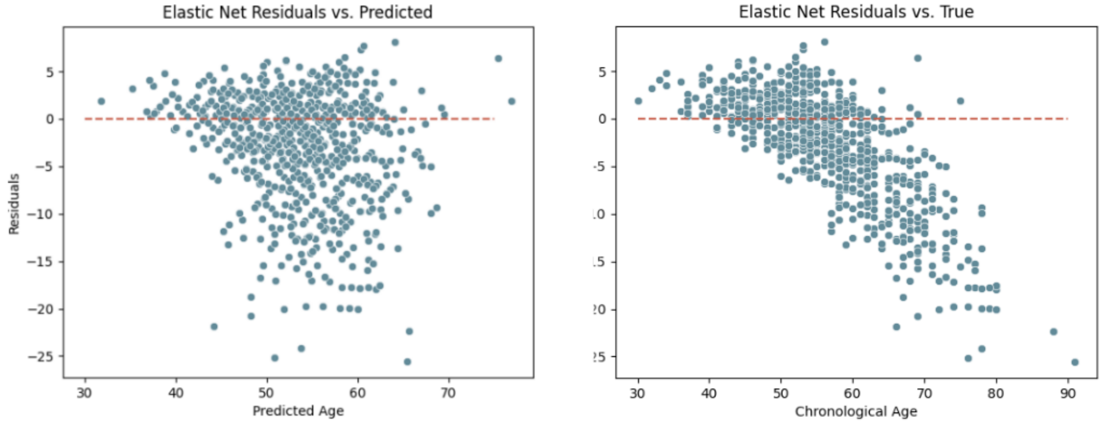


Figure 2: Plots of residuals versus predicted and true age from the ElasticNet model. There is a clear negative trend in the residuals versus chronological age, indicating a shrinkage pattern for ElasticNet.

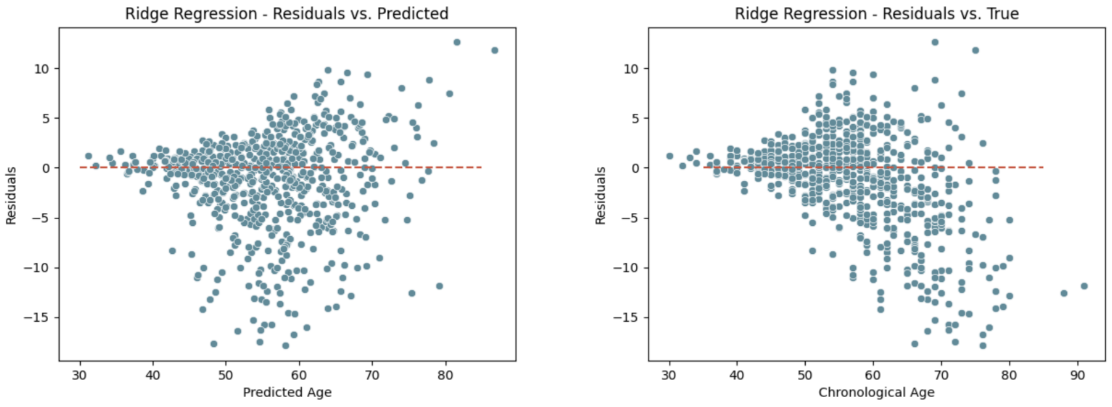


Figure 3: Plots of residuals versus predicted and true age from the ridge regression model. Plots demonstrate improved performance of the model compared to ElasticNet with residuals evenly distributed across both predicted and chronological ages. Smaller MAE and residual error is to be expected for lower ages in the training cohort due to age distribution matching.

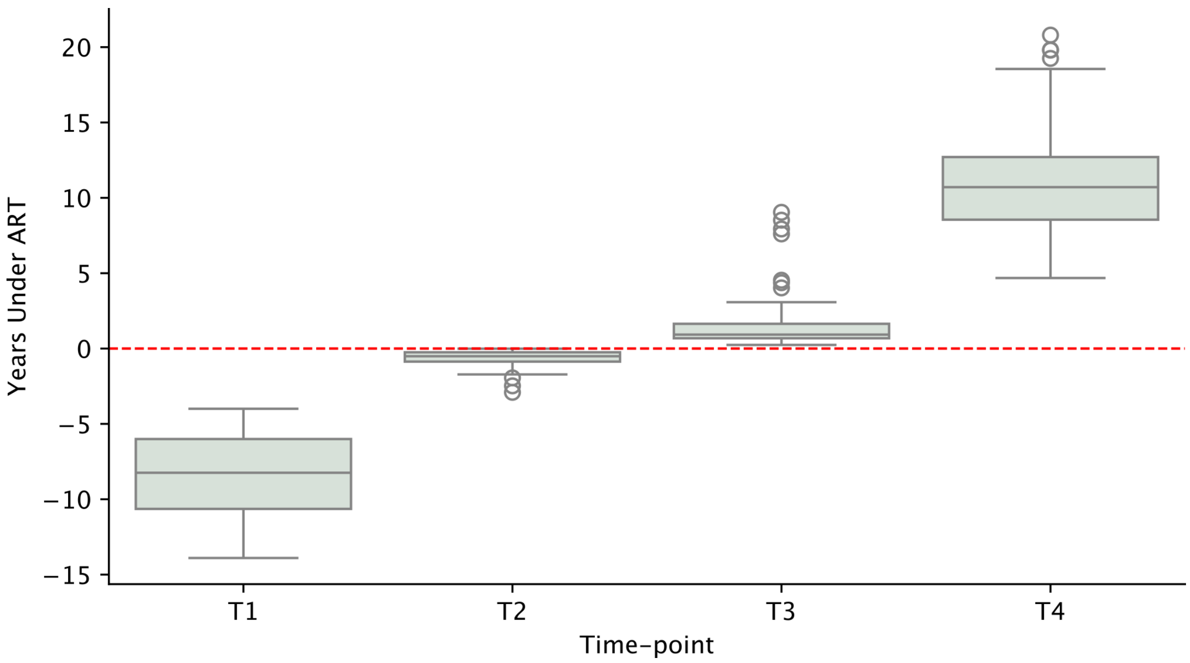


Figure 4: Duration of ART in longitudinal test cohort split across the fours cross-sectional time-points

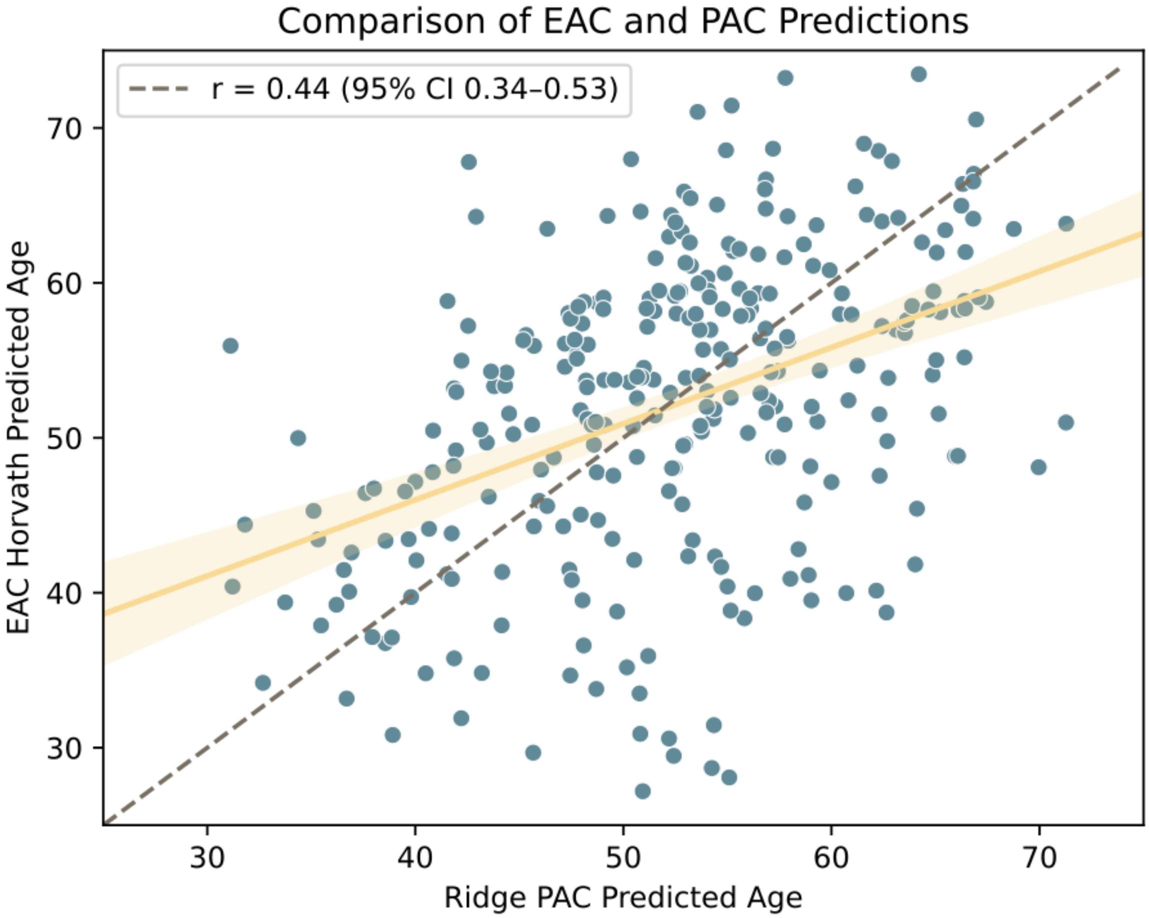


Figure 5: Moderate correlation between PAC and EAC demonstrate shared directionality and behaviour of predictions.

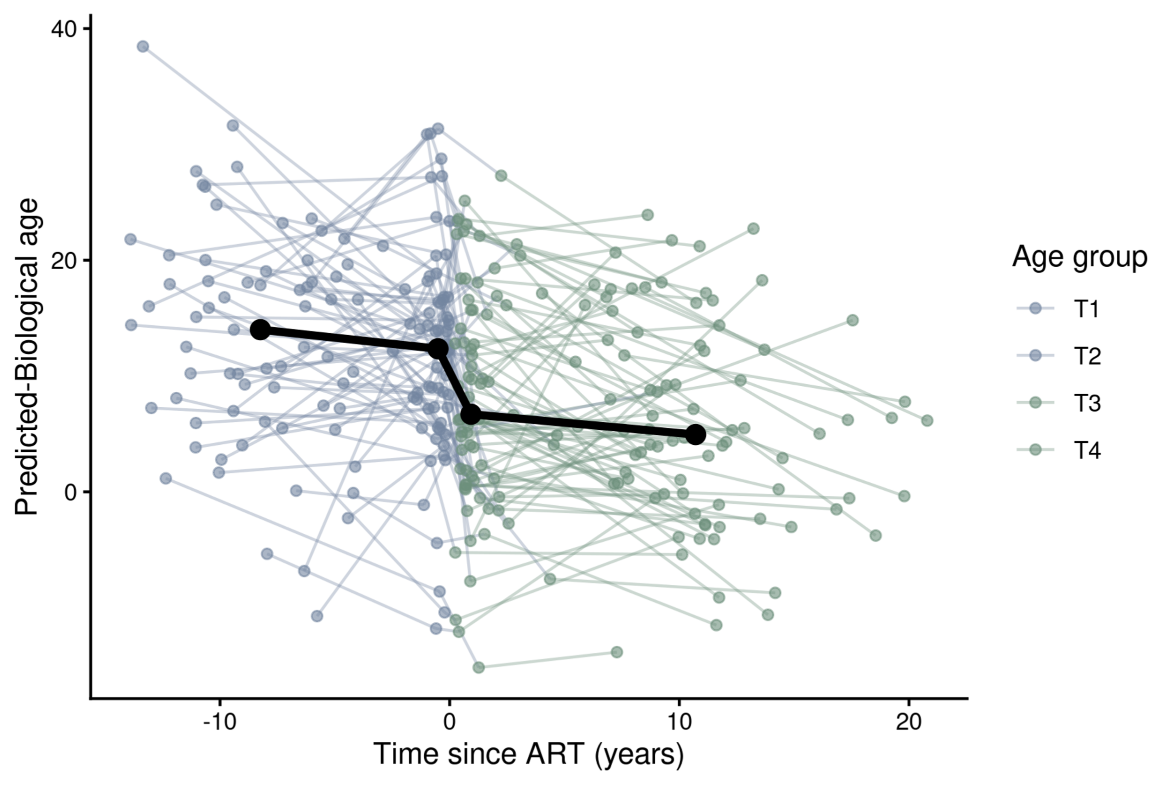


Figure 6: Individual trajectories plot with median trajectory between time-points

#### 12 Supplementary Tables

Table 1: Age-based loss and distribution matching metrics in training cohort for linear ridge-regression model.

| Age Bin | n | MAE | RMSE | Mean Age per Bin |
|---|---|---|---|---|
| (29.999, 48.0] | 150.0 | 0.965904 | 1.226247 | 43.973333 |
| (48.0, 54.0] | 168.0 | 2.238194 | 2.957395 | 51.910714 |
| (54.0, 58.0] | 121.0 | 2.978583 | 3.789012 | 56.636364 |
| (58.0, 65.0] | 148.0 | 3.765820 | 4.839222 | 61.547297 |
| (65.0, 91.0] | 140.0 | 7.042657 | 8.486538 | 70.664286 |

Table 2: Age-based loss and distribution matching metrics in test cohort at T4 time-point for linear ridge-regression model.

| Age Bin | n | MAE | RMSE | Mean Age per Bin |
|---|---|---|---|---|
| (20.999, 37.0] | 65.0 | 13.950335 | 16.193773 | 32.107692 |
| (37.0, 43.0] | 60.0 | 10.543550 | 12.705343 | 40.533333 |
| (43.0, 47.0] | 52.0 | 9.580119 | 11.338162 | 45.480769 |
| (47.0, 54.0] | 61.0 | 5.908254 | 7.723658 | 50.901639 |
| (54.0, 74.0] | 56.0 | 6.425241 | 7.690319 | 61.000000 |

Table 3: OLS inference for the effect of ART initiation on PAA.

| | Coefficient | Std. error | z | 95% CI |
|---|---|---|---|---|
| Intercept | 10.31 | 1.01 | 10.22 | [8.34, 12.29] |
| Post-ART (vs Pre-ART) | -5.99 | 0.89 | -6.76 | [-7.72, -4.25] |

Table 4: OLS inference for the effect of ART initiation on PAA compared to the first baseline time-point T1.

| | Coefficient | Std. error | z | P-value | 95% CI |
|---|---|---|---|---|---|
| Intercept | 10.71 | 1.15 | 9.30 | <0.001 | [8.46, 12.97] |
| T2 (vs T1) | -0.80 | 1.10 | -0.73 | 0.466 | [-2.96, 1.36] |
| T3 (vs T1) | -4.88 | 1.09 | -4.48 | <0.001 | [-7.02, -2.75] |
| T4 (vs T1) | -7.90 | 1.13 | -7.00 | <0.001 | [-10.11, -5.68] |

Table 5: OLS inference for the effect of covariates on proteomic age predictions

| | Coefficient | Std. error | z | P-value | 95% CI |
|---|---|---|---|---|---|
| Intercept | -0.9441 | 0.411 | -2.298 | 0.022 | [-1.749, -0.139] |
| Center 1 | -1.2145 | 0.633 | -1.919 | 0.055 | [-2.455, 0.026] |
| Center 2 | -0.6428 | 0.491 | -1.311 | 0.190 | [-1.604, 0.319] |
| Center 3 | -0.7703 | 0.583 | -1.321 | 0.187 | [-1.913, 0.373] |
| Center 4 | -0.5714 | 0.671 | -0.851 | 0.395 | [-1.887, 0.744] |
| Center 5 | -1.2111 | 0.951 | -1.273 | 0.203 | [-3.076, 0.653] |
| Center 6 | -0.1114 | 0.570 | -0.196 | 0.845 | [-1.229, 1.006] |
| Sex | 2.2845 | 0.466 | 4.899 | 0.000 | [ 1.371, 3.198] |
| Genetic PC1 | 1.228e-05 | 1.13e-05 | 1.091 | 0.275 | [-9.78e-06, 3.44e-05] |
| Genetic PC2 | 117.5462 | 19.695 | 5.968 | 0.000 | [78.945, 156.147] |
| Genetic PC3 | 31.6245 | 22.296 | 1.418 | 0.156 | [-12.075, 75.324] |

Table 6: OLS inference for the effect of cardiac case screen on proteomic age predictions

| | Coefficient | Std. error | z | P-value | 95% CI |
|---|---|---|---|---|---|
| Intercept | -1.2798 | 0.249 | -5.135 | 0.000 | [-1.768, -0.791] |
| Cardiac Case (c=1) | 0.1887 | 0.348 | 0.542 | 0.588 | [-0.494, 0.871] |

Table 7: PAC vs. EAC comparison of advanced ageing predictions at each cross-sectional time-point

| Time-point | n | Mean PAA | Mean EAA | Mean PAA - EAA | Std PAA - EAA | T | p-value | CI95 Low | CI95 High |
|---|---|---|---|---|---|---|---|---|---|
| T1 | 67 | 10.71 | 6.75 | 3.96 | 10.84 | 2.99 | 0.00 | 1.32 | 6.61 |
| T2 | 67 | 9.91 | 10.61 | -0.70 | 9.66 | -0.59 | 0.55 | -3.06 | 1.65 |
| T3 | 80 | 5.83 | 7.66 | -1.83 | 8.79 | -1.86 | 0.07 | -3.79 | 0.13 |
| T4 | 80 | 2.82 | 2.93 | -0.12 | 9.28 | -0.11 | 0.91 | -2.18 | 1.95 |
